# Detection of SARS-CoV-2 RNA on public surfaces in a densely populated urban area of Brazil

**DOI:** 10.1101/2020.05.07.20094631

**Authors:** Jônatas Santos Abrahão, Lívia Sacchetto, Izabela Mauricio Rezende, Rodrigo Rodrigues, Ana Paula Correia Crispim, César Moura, Diogo Correa Mendonça, Erik Reis, Fernanda Souza, Gabriela Fernanda Garcia Oliveira, Iago José da Silva Domingos, Paulo Boratto, Pedro Henrique Bastos e Silva, Victoria Fulgêncio Queiroz, Thaís Bárbara de Souza Silva, Graziele Pereira Oliveira, Viviane de Souza Alves, Pedro Augusto Alves, Erna Geessien Kroon, Giliane de Souza Trindade, Betânia Paiva Drumond

**Affiliations:** Universidade Federal de Minas Gerais, Departamento de Microbiologia, Laboratório de Vírus; Universidade Federal de Ouro Preto, Departamento de Ciências Biológicas, Laboratório de Biologia e Tecnologia de Microrganismos; Universidade Federal de Minas Gerais, Departamento de Ecologia, Genética e Evolução, Laboratório de Genética Molecular de Protozoários Parasitas; Instituto René Rachou – FIOCRUZ, Laboratório de Imunologia de Doenças Virais - IDV; Universidade Federal de Minas Gerais, Departamento de Microbiologia, Laboratório de Biologia Celular de Microrganismos

## Abstract

**Importance:** The COVID-19 pandemic has resulted in more than 3.5 million cases and 245 thousand deaths worldwide as of May 6, 2020. Determining the extent of the presence of the virus on public surfaces is critical for understanding the potential risk of infection in these areas.

**Objective:** To evaluate the presence of SARS-CoV-2 RNA on public surfaces in a densely populated urban area in Brazil.

**Design and Setting:** A total of 101 samples were collected from different surfaces in public places in the region of Belo Horizonte with the highest number of COVID-19 cases. Samples were collected near the hospital and public transportation areas using sterile swabs, and then submitted to nucleic acid extraction and genomic detection and quantification by one-step qPCR.

**Results:** Seventeen of the 101 samples tested positive (16.8%) for SARS-CoV-2 RNA, including samples from bus stations/terminals, public squares, and sidewalks, including those near hospitals.

**Conclusions and Relevance:** Our data indicated the contamination of public surfaces by SARS-CoV-2, especially near hospital areas, highlighting the risk of infection for the population. Constant monitoring of the virus in urban areas is required as a strategy to fight the pandemic and prevent further infections.

**Key points:** 

**Question:** 

Can SARS-CoV-2 be detected on surfaces in public areas used by a large proportion of the population?

**Findings:** SARS-CoV-2 RNA was detected in different locations, including bus stations/terminals, squares, and sidewalks, especially those near hospitals, in a densely populated area of Belo Horizonte, Brazil.

**Meaning:** This study highlights the need for constant monitoring for the presence of SARS-CoV-2 RNA in urban areas to support better strategies to fight the COVID-19 pandemic and prevent further infections.

## Text

Since late December 2019, the world has experienced the worst global health crisis in recent decades. This pandemic, called coronavirus disease 2019 (COVID-19), is caused by severe acute respiratory syndrome coronavirus 2 (SARS-CoV-2) and has affected several sectors, including those related to medicine, economics, and politics, among others.^1–4^ COVID-19 has already affected over 200 countries, resulting in more than 3.5 million cases and 245 thousand deaths worldwide as of May 6, 2020.^5^ In Brazil, the first official case was registered on February 26, 2020, and more than 126,000 confirmed cases and 8,500 deaths had been registered across the country as of May 6, 2020.

Belo Horizonte is the capital of Minas Gerais State, one of the most populous metropolitan regions in Brazil (a population of 6 million people). The city recorded 845 confirmed cases and 20 deaths due to COVID-19.^6^ The areas within Belo Horizonte where most of the deaths and confirmed cases have occurred correspond precisely to areas with public squares, bus stations/terminals, and hospital areas, i.e., where a large flow and concentration of people is commonly observed.^6^

Recent studies have identified the presence of SARS-CoV-2 RNA on different surfaces and environments inside hospitals, revealing the dynamics of viral dissemination within these places.^7,8^ SARS-CoV-2 remains viable on different types of surfaces, such as metal and plastic, for up to 72 hours, depending on the type of surface material, and can remain infectious in aerosols for at least 3 hours.^9^ Assessing the presence of the virus in the environment, objects, and surfaces in public areas is fundamental for understanding the risk of infection in the population. In addition, any information gained can be used by health managers to control population movements in these areas, as well as implement environmental disinfection measures.

In this context, we investigated the presence of SARS-CoV-2 RNA in a downtown area of Belo Horizonte in April 2020 (Figure 1). This region of the city (central-southeast) has the highest concentration of hospitals and health units and the highest number of notified COVID-19 cases (Figure 1). Importantly, this part of the city also has a large number of people accessing public transportation and transportation facilities daily. A total of 101 samples were collected from the following 30 locations (Figure 1): i) 15 bus stations (stops) (for each bus station, one sample was collected from a bench and two from the ground just below the bench. Total = 45 samples); ii) front-door sidewalk of eight hospitals (1–3 collections per location. Total = 12 samples); iii) four bus terminals (entrance handrails) (6 samples per terminal. Total = 24); iv) three benches and tables in public squares (total = 20 samples). For the collection of environmental samples, swabs with sterile phosphate-buffered saline were vigorously rubbed on surfaces (10 cm^2^) of the aforementioned objects. The swabs were then transferred to tubes containing transport solution (1 mL of guanidine isothiocyanate buffer, 4 M). For each sample, 70 of transport solution containing a sample was submitted to nucleic acid extraction using the QIAmp Viral RNA Mini Kit (QIAGEN, Maryland, USA). Total RNA (5 μL) was used as a template for one-step qPCR (Promega, Wisconsin, USA) (in a final volume of 20 μL per reaction, GoTaq1-sept qPCR system, Promega), using primers and probes specific for the N1 and N2 regions of the SARS-CoV-2 genome (CDC, USA, 2020). Samples were considered positive if both targets had a cycle threshold (Ct) less than 40 (CDC, USA, 2020). Negative (extraction control and non-template control) and positive controls (RNA extracted from inactivated SARS-CoV-2, kindly provided by Dr. Danielle Durigon and Dr. Edison Durigon, USP, Brazil) were used. To confirm the results, all positive samples were submitted to a second round of RNA extraction and RT-qPCR. Quantification of viral RNA in the environmental samples was based on a standard curve generated from serial dilutions (1:10) of SARS-CoV-2 RNA and converted to genomic units per square meter of surface (N1 gene). SARS-CoV-2 RNA control was previously quantified as described elsewhere.^10^

**Fig 1.**
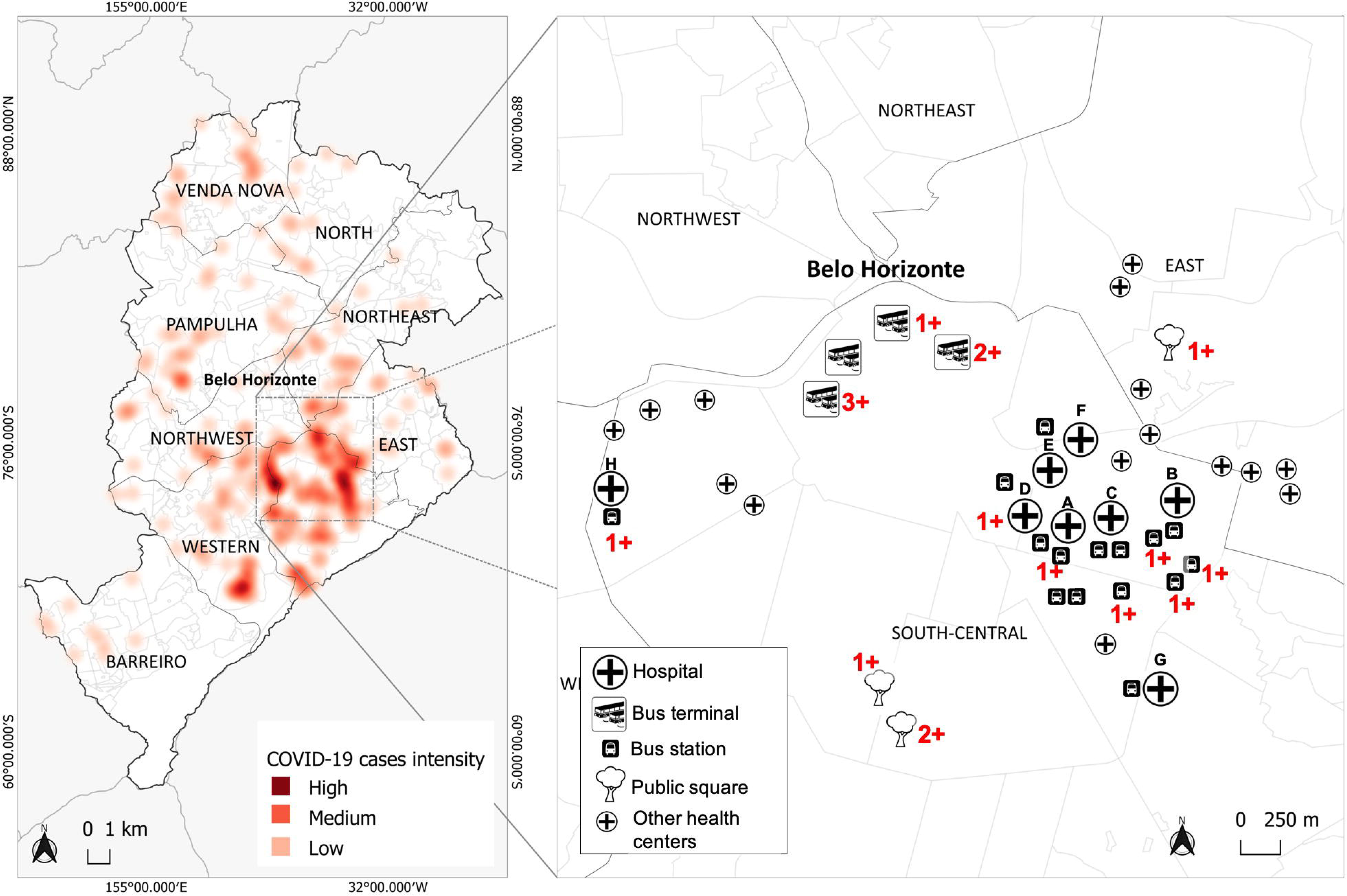
Map of Belo Horizonte showing the areas with positive cases of COVID-19 and collection sites. **Left:** red points on the map indicate the occurrence of positive cases reported by the City Hall as of May 4, 2020. The color intensity indicates the number of reported cases. **Right:** downtown area of Belo Horizonte where the samples were collected. Hospitals, bus stations (stop), bus terminals and public squares enrolled in this study are indicated (see legend). In this map we also depicted other health centers (small crosses) in this studied area. Numbers in red indicate the location and the quantity of positive samples detected per site.

A total of seventeen samples (16.8%) were positive for the presence of SARS-CoV-2 RNA (Table 1). SARS-CoV-2 RNA was detected in 6 of the 15 tested bus stations (5.9%) located in front of hospitals, with loads ranging from 20 to 620 genomic units/m^2^. Four of these positive samples were collected from bus station benches and two from the ground under the benches. The following hospitals presented SARS-CoV-2-positive bus stations: hospital A, 1 positive/4 assayed stations; hospital B (3 positive/4 assayed stations); hospital C (1 positive/3 assayed stations); and hospital H (1 positive/1 assayed station). We also detected SARS-CoV-2 RNA in the front-door sidewalk of hospital D (0.9%) (2,990 genomic units/m^2^). SARS-CoV-2 RNA was detected in three of the four bus terminals tested, in a total of six handrails (5.9%), three from terminal 1, one from terminal 3, and two from terminal 4 (50 to 2,110 genomic units/m^2^). Finally, we detected viral RNA in four samples from three public squares: one from a concrete table in square A, and three from concrete benches – one from square B and two from square C (3.9%) (20 to 200 genomic units/m^2^). The Belo Horizonte City Hall was informed about the contaminated areas, and, after disinfection (laundry detergent followed by 1% sodium hypochlorite), viral RNA could no longer be detected.

**Table 1.** | RNA concentration of SARS-CoV-2 in surfaces at different locations in Belo Horizonte^#^

| Category | Sites | Surface | Concentration (genomic units/m<sup>2</sup>) |
| --- | --- | --- | --- |
| **Hospitals` bus stations (BS)** |  |  |  |
|  | Hospital A BS2 | Bench (metal) | 620 |
|  | Hospital B BS2 | Bench (metal) | 80 |
|  | Hospital B BS3 | Bench (metal) | 70 |
|  | Hospital B BS1 | Ground (concrete) | 20 |
|  | Hospital C BS1 | Ground (concrete) | 60 |
|  | Hospital H BS1 | Bench (metal) | 150 |
| **Hospitals` front door ground (FDG)** |  |  |  |
|  | Hospital D FDG1 | Ground (concrete) | 2990 |
| **Bus terminal handrails (Hr)** |  |  |  |
|  | Bus terminal 1 Hr1 | Handrail (metal) | 200 |
|  | Bus terminal 1 Hr2 | Handrail (metal) | 50 |
|  | Bus terminal 1 Hr3 | Handrail (metal) | 2110 |
|  | Bus terminal 3 Hr5 | Handrail (metal) | 520 |
|  | Bus terminal 4 Hr1 | Handrail (metal) | 70 |
|  | Bus terminal 4 Hr5 | Handrail (metal) | 2080 |
| **Public squares** |  |  |  |
|  | Public square 1 | Table (concrete) | 200 |
|  | Public square 2 | Bench (concrete) | 20 |
|  | Public square 3 | Bench (concrete) | 70 |
|  | Public square 3 | Bench (concrete) | 70 |

<sup>#</sup> Only positive samples are listed in this table. For more information about collected samples and locations, please see Figure 1 and Sup. Material.

Although we sought to detect the presence of viral RNA, not infectious particles, it is highly plausible that infectious particles were also present in these environments, and care must be taken to avoid further contamination and the eventual collapse of the local health system. Despite a relatively small number of samples, it is surprising that almost 20% of the sampled sites were positive for the presence of the virus, especially when dealing with places with a large flow of pedestrians.

Although Belo Horizonte has relatively few cases when compared with other cities in Brazil, such as São Paulo and Rio de Janeiro, our data indicate that the virus is circulating in the city and can be found on surfaces such as benches, tables, handrails, and floors, and in places with a large flow of people, such as public squares and hospital entrances. The detection of the viral RNA at these sites indicates that adequate cleaning of public environments and reinforcement of educational campaigns for hygienic and social distancing practices should be undertaken.

Our study highlights the need for the constant assessment of the presence of the virus, not only in hospital facilities, but also in places close to medical areas and with a large circulation of people. The presence of SARS-CoV-2 in these environments can result in an increase in the number of cases of the disease in the near future if control measures are not forcefully adopted.

## Data Availability

All data presented in this manuscript is available.

## Authors’ contributions

Design of the study: J.S.A., B.P.D., G.S.T., E.G.K., V.G.; collection of samples: J.S.A.; RNA extraction, qPCR and analyses: L.S., I.M.R., R.R., A.P.C.C., C.M., D.C.M., E.R., F.S., G.F.G.O., I.J.S.D., P.B., P.H.B.S., V.F.Q., T.B.S.S., G.P.O., P.A.A.; Manuscript writing: R.R., J.S.A, E.G.K., G.S.T., B.P.D., V.S.A.. All authors have read and approved the final version of the manuscript.

## Acknowledgments

We thank our colleagues from Laboratório de Vírus – UFMG (Universidade Federal de Minas Gerais) for their assistance. In addition, we thank the Pró Reitorias de Pesquisa da UFMG/Secretaria de Educação Superior/Ministério da Educação (number 23072.211119/2020-10), Programa de Pós-graduação em Microbiologia da UFMG, CNPq (Conselho Nacional de Desenvolvimento Científico e Tecnológico), CAPES (Coordenação de Aperfeiçoamento de Pessoal de Nível Superior) and FAPEMIG (Fundação de Amparo à Pesquisa do estado de Minas Gerais) for their financial support. B.P.D, E.G.K., G.S.T. and J.S.A. are a CNPq researchers. J.S.A. and P.A.A. are members of Rede Vírus MCTIC. We also thank Belo Horizonte City Hall for its assistance for contaminated areas disinfection.

